# Experiences of COVID-19 Recovered Patients – A Qualitative Case Study from a Hotspot in Saudi Arabia

**DOI:** 10.1101/2021.03.01.21252508

**Authors:** Abdulrahman Alhajjaji, Ahmad Kurdi, Sultan Faqeh, Safwan Alansari, Akrm Abdulaziz, Moayad Allihyani, Omar Almaghamsi, Ejaz Cheema, Majid Ali

## Abstract

**Background:** COVID-19 is highly contagious and can have fatal outcomes in the elderly and those with comorbidities. Social distancing is highly recommended by the World Health Organization to prevent the spread of the disease. However, it is difficult to maintain social distancing in highly populated areas where people live in close proximity. Such high-risk areas have the potential to become hotspots for the disease spread, should one person therein contract the disease. Nakkasah is one such area in the Makkah city of Saudi Arabia which has been a hotspot in this pandemic. This study aims to qualitatively explore the experiences of COVID-19 recovered patients residing in this area.

**Methods:** We employed semi-structured face-to-face interviews with people living in Nakkasah, above 18 years of age, and recovered from COVID-19. An interview guide was developed, validated, piloted, and minor changes were made. Two trained students conducted the interviews in the Arabic language in a semi-private area of the community center. The interviews were audio-recorded, with informed consent from interviewees, transcribed verbatim, and thematically analyzed later.

**Results:** Eleven eligible COVID-19 recovered people (two female and nine male) agreed to be interviewed, and their verbal informed consent was audio recorded. The mean interview time was 24 minutes. Thematic analysis generated 30 subthemes, which were categorized into seven overarching themes: information about COVID-19; life during COVID-19 illness; spreading of COVID-19; precautionary measures; interventions that helped in recovery; impact of COVID-19 on life; support received during COVID-19 illness.

**Conclusion:** Experiences of people from the hotspot who had recovered from COVID-19 highlighted how life had been like in the hotspot under lockdown especially with having been afflicted with the infection, factors that facilitated their recovery, and the way their lives were and have been affected due to COVID-19.

## BACKGROUND

The coronavirus disease 2019 (COVID-19) pandemic is essentially a global health crisis. It has caused significant health-related morbidity and mortality around the world.^1^ The disease is highly contagious and leads to fatal outcomes in specific populations, especially those with comorbidities as well as the elderly.^2^ According to the World Health Organization (WHO), the current evidence suggests that the disease spreads between people by two main methods, directly (through close contact with an infected person via the mouth or nasal secretions) or indirectly (through contaminated objects or surfaces).^3^ The WHO also states that people who are in close contact (within one meter) with an infected person can catch COVID-19 when the infectious droplets enter their mouth, nose, or eyes. The WHO has put forward several recommendations to prevent the spread directly or indirectly. One of these recommendations is to maintain social distancing which is currently defined as keeping a distance of at least one meter from others in open public places such as parks and walking areas as well as in the confined places such as shops, restaurants, etc. The implementation of social distancing is undoubtedly challenging in densely populated or crowded areas where people live in close proximity. People living in such areas are at increased risk of contracting the infection due to the rapid transmission of the disease.

Nakkasah is one of such thickly populated areas of Makkah city in the Kingdom of Saudi Arabia (KSA). The houses and residential buildings are very congested with narrow streets and poor sanitation. Most of the population comprises migrants from Burma with some Africans, Bangladeshis, and Indonesians and mainly represent the lower socio-economic class. The predominant language is Burmese.^4^ The living conditions in the area pose a high health risk to the residents. The area had been declared a hotspot in the current COVID-19 pandemic due to the rapid and uncontrolled transmission of the infection both within the local community as well as the spread of the disease to the other areas of the city. Consequently, the area was put under complete lockdown for a period of three months (April-June 2020) and was served by volunteer healthcare organizations and KSA’s Ministry of Health teams. A local community committee was responsible for overseeing the local social affairs and employed a Saudi translator to facilitate the communication between the local community and the Ministry of Health teams.

It is paramount to explore the experiences of COVID-infected patients from the time of their diagnosis or the emergence of their symptoms through to their recovery, especially the patients from hotspots. This can not only help the healthcare organizations to adopt new care models and streamline the workflows but also help improve the quality of life of infected patients in high-risk communities,^5^ during the current and future outbreak of pandemic diseases. To the authors’ knowledge, there is currently no published study that has qualitatively explored the experiences of COVID-19 recovered patients living in a hotspot area. This study, therefore, aimed to qualitatively explore the experiences of COVID-19 recovered patients in such an area to provide a useful insight into how the quality of care and life can be improved for patients in high-risk communities such as Nakkasah during this pandemic and in future disease outbreaks.

## METHODS

This study was approved by the biomedical ethical committee of Umm Al-Qura University, Makkah, KSA.

### Study design and sampling

In this study, we adopted qualitative methodology and employed semi-structured face-to-face interviews with the eligible participants who were recruited on a convenience sampling basis. The official translator and volunteers working in Nakkasah helped identify the potential participants for the interviews.

### Participants and setting

The participants living in Nakkasah, above 18 years of age, and having suffered and recovered from COVID-19 during the lockdown period (either hospitalized or not during their illness), were deemed eligible for the recruitment. Prior to the recruitment of the participants, formal written permission was sought from the community leader of Nakkasah. The interviews were conducted in a semi-private area of the community center. No financial incentive was offered to the participants.

### Semi-structured interviews

To facilitate the interviews, the interview guide was developed in the English language and then translated into the Arabic language by the authors. A bilingual academic staff member double-checked the accuracy of the translation. The interview guide comprised open-ended questions related to six topics (Panel 1). The interview guide was checked for face validity with two experienced academic staff members and piloted with two recovered patients. Minor changes were made in the interview guide following the validation and piloting. The interviews were expected to take 15-20 minutes.

Two research students were trained to conduct the semi-structured interviews. The interviews were conducted in the Arabic language with the recommended health precautionary measures in place. Informed consent was taken from each participant prior to the interview. The interviews were audio-recorded and transcribed verbatim later. The accuracy of the transcriptions was checked alternatively by the authors.

### Data analysis

Qualitative data in the transcriptions were analyzed using the inductive method of thematic analysis. The initial step of the analysis involved familiarizing with the data through reading the transcripts. It was followed by a manual generation of initial codes from the data by the two teams of authors independently. The coding was then reviewed and verified by the academic supervisors. This was followed by the categorization of codes into potential subthemes and themes by the two teams of authors independently, which was further reviewed and verified by the academic supervisors. Any discrepancies and inaccuracies at all steps were resolved through discussion.

## RESULTS

We interviewed 12 eligible participants in August 2020. One interview (Interview 8) was excluded as the participant did not attend the interview in person and instead sent the representative. Therefore, 11 interviews (two female and nine male participants) were included in the analysis (Table 1). Data saturation was achieved with ten interviews; however, the 11^th^ interview was included in the analysis to confirm the data saturation. Six participants could not speak the Arabic language. The interviews with these participants were conducted with the help of the official translator of the Nakkasah community. Thematic analysis generated 378 codes which were categorized into 30 subthemes. These subthemes were further categorized into seven overarching themes (Panel 2 and Table 2).

**Table 1.** Characteristics of the participants

| **Participant** | **Gender** | **Age range (years)** | **Presence of translator in the interview** |
| --- | --- | --- | --- |
| Participant 1 (P1) | Female | 21-30 | No |
| Participant 2 (P2) | Male | 11-20 | No |
| Participant 3 (P3) | Male | 51-60 | Yes |
| Participant 4 (P4) | Male | 51-60 | Yes |
| Participant 5 (P5) | Male | 21-30 | No |
| Participant 6 (P6) | Male | 41-50 | No |
| Participant 7 (P7) | Male | 61-70 | Yes |
| Participant 9 (P9) | Male | 41-50 | Yes |
| Participant 10 (P10) | Male | 41-50 | Yes |
| Participant 11 (P11) | Female | 71-80 | Yes |
| Participant 12 (P12) | Male | 31-40 | No |

**Table 2.** Themes with the number of associated subthemes and codes

| **Themes** | **Number of associated subthemes** | **Number of associated codes** |
| --- | --- | --- |
| 1. Information about COVID-19 | 3 | 43 |
| 2. Life during COVID-19 illness | 8 | 76 |
| 3. Spreading of COVID-19 | 2 | 21 |
| 4. Precautionary measures | 2 | 46 |
| 5. Interventions that helped in recovery | 4 | 29 |
| 6. Impact of COVID-19 on life | 5 | 84 |
| 7. Support received during COVID-19 illness | 6 | 79 |

### 1. Information about COVID-19

When asked about the sources of information for COVID-19, the majority of the participants mentioned television or social media applications such as WhatsApp, Twitter, YouTube, Snapchat and Instagram.

Interviewee: “*Photos were sent to me by WhatsApp, (and also) the audios sent by people to spread the awareness*.” (Interview 1)

These participants reported that the disease is spread by touching the contaminated surfaces, shaking hands, transferring paper money and coins from hand to hand, not wearing gloves, and by social gatherings. They also mentioned that children, adolescents, older people and those with chronic disease (such as respiratory problems) are considered to be high-risk patients.

Others were unaware of how the virus spreads or what precautionary measures to observe as they did not seem to be using social media.

Translator: “*He says, I don’t know because I don’t follow the news on television or social media*.” (Interview 10)

### 2. Life during COVID-19 illness

The participants explained how their life was with the illness during the lockdown period in Nakkasah. The majority of the participants reported being hospitalized during the illness. One participant described how the transition of care affected the mental status.

Interviewee: “*I suffered (with COVID-19) for a long time, so when I came back home (from the hospital), they took me back to the hospital the next day and I stayed (there) for an even longer period. They said that my psychological condition was very bad and after that, they took me to the hotel (designated for quarantine)*.” (Interview 1)

The participants reported being quarantined for the average duration of 13-18 days. The majority of them were quarantined in designated hotels. Most of the participants suffered from cough and fever and some of them reported these symptoms as the first sign of the infection. Shortness of breath and chest tightness were also reported as the initial symptoms by a few participants. Some of the participants mentioned that they also suffered from a headache that prevented them from walking. Some participants reported unusual long-term symptoms that were not directly related to COVID-19.

Interviewee: “*I was not having a hair loss in the past but now I am having hair loss (due to COVID-19)*.” (Interview 1)

The participants also described their concerns and worries during the quarantine.

Translator: “*He said, I felt I was close to my death*.” (Interview 9)

Translator: “*He said, when someone is in the hospital or hotel (due to COVID-19), no one visits him*.” (Interview 11)

The majority of the participants deciphered that they had recovered from COVID-19 owing to the relieving of the symptoms or the negative COVID-19 test. Diabetes and hypertension were reported to be the most common comorbidities by the majority of the participants. Some participants reported that their comorbid conditions were not affected by COVID-19 illness whereas others mentioned the worsening of their comorbid conditions during COVID-19 illness along with “*very bad*” COVID-19 symptoms. One participant linked the worsening of comorbid conditions to the reduced availability of regular medications.

Interviewee: “*I have diabetes, hypertension and heartburn and I felt very bad (due to these conditions) because I could not get the medications like before*.” (Interview 1)

### 3. Spreading of COVID-19

Some of the participants mentioned that they might have caught the infection through close contact with the infected family members or friends, whereas others thought that they might have contracted the virus from asymptomatic infected people around them.

Interviewee: “*Well, I went to meet my relatives (who were infected) and I touched the contaminated surfaces there like doors*.” (Interview 2)

Interviewee: “*I got the infection from my brother who goes out to get grocery, but he did not have any symptoms and did not look sick*.” (Interview 1)

The majority of the participants believed that they did not spread the infection to others, whereas a few of them thought that they might have spread it to their relatives due to close contact.

Interviewee: “*I isolated myself and didn’t spread it to anyone*.” (Interview 9)

Interviewee: “*My son who was responsible for taking me to the hospital was fine and had no symptoms, and then he got infected because he was accompanying me*.” (Interview 10)

### 4. Precautionary measures

The WHO has recommended precautionary measures to prevent the spread of COVID-19. Several of these measures were mentioned and observed by the participants during the lockdown such as washing hands frequently, social distancing and wearing masks and gloves.

Interviewee: “*We were trying to avoid touching anything, wearing gloves and masks and applying social distancing by keeping a distance of one to two meters from people*.” (Interview 1).

Some participants reported that they were still observing some of the precautionary measures even after the lockdown was lifted such as avoiding gatherings and maintaining social distancing, however, to the lesser extent as compared to the precautionary measures observed during the lockdown.

Interviewee: “*Gatherings are now reduced, we wash hands, etc. but not as much as during the lockdown*.” (Interview 1).

### 5. Interventions that helped in recovery

The majority of the participants reported that they used the medicines to control the symptoms only and not for the cure.

Interviewee: “*Effervescent form of (paracetamol brand mentioned) helped me to reduce my symptoms but did not treat me*.” (Interview 2)

Some participants mentioned that they were also prescribed antibiotics and anti-allergy medications for COVID-19. The participants also reported that they were provided with information regarding the reason for prescribing these medicines and how to use them. Other participants, on the appearance of initial symptoms, took home remedies such as honey, lemon, and ginger to relieve the symptoms. One participant reported using these remedies because of the fear of seeing the doctor.

Translator: “*He says, in the beginning, he didn’t want to go to the doctor. He said, he was afraid of seeing the doctors, so he used natural (remedies)*.” (Interview 9)

Some participants stated that they took medicines as well as home remedies.

### 6. Impact of COVID-19 on life

Since Nakkasah was the only area under complete and strict lockdown in KSA, people living herein had a unique experience. The majority of the participants showed concerns on hearing the news of Nakkasah being under lockdown.

Interviewee: “*I (was afraid to) go outside from that day*.” (Interview 2)

One participant described his situation of being referred to the psychiatrist due to mental stress related to the quarantine as well as the concern regarding the community.

Interviewee: “*This is one of the psychological symptoms that affected me, because they said that they will move Nakkasah (to outside the city due to the spread of COVID-19)*.” (Interview 1)

Some participants indicated the effect of lockdown on life.

Interviewee: *“(Social gatherings) are reduced now, not like always*.” (Interview 11 – Line 305).

The financial status of the people in Nakkasah was reported to be adversely affected during the lockdown.

Translator: “*She felt sad because the people in Nakkasah go out for their daily sustenance and their financial status was hard (to maintain during the lockdown)*.” (Interview 11)

Some participants reported lingering side effects after the recovery from COVID-19, such as irritable bowel syndrome, hair loss, and shortness of breath.

Interviewee: “*My colon does not feel the same (after recovery)*.” (Interview 12).

Interviewee: “*I did not have shortness of breath and hair loss before and now I’m being treated for these symptoms*.” (Interview 1)

### 7. Support received during COVID-19 illness

The majority of the participants indicated and acknowledged the emotional support such as phone calls and daily messages they received from family and friends while they were quarantined during the lockdown period. The participants also highlighted the support received from the government (Ministry of Health) and the community committee in Nakkasah.

Interviewee: “*My friends and family always used to call me and care about me, and I never felt alone (because) they asked and cared*.” (Interview 1)

One participant was overwhelmed with emotions while describing the support received from the government.

Translator: “*She said that she cannot describe (the feelings) [participant crying] and doesn’t know how to thank and pray for the (KSA’s) king and Ministry of Health*.” (Interview 11)

The community committee supported by supplying food and ensured that the people were observing precautionary measures.

Interviewee: “*The community provided us (with) food supplies*.” (Interview 9)

## DISCUSSION

COVID-19 lockdowns can paralyze the functioning society. However, the individuals and especially those who suffer from COVID-19 continue with their life with its basic needs. Exploring their experiences provides an insight into how their lives are affected and measures that can be taken, on individual and social levels, to ease their lives and make them valuable members of society again.

This study indicates that the participants who were social media users were well-aware of the COVID-19 and the related issues. Approximately 70% of the population in KSA actively use social media.^6^ Due to the widespread use of social media in KSA, we expect reasonably good awareness regarding COVID-19 in society. This coincides with the findings of the survey conducted in KSA to determine the COVID-19 awareness, which reported moderate to high (60-80%) awareness in the society with the Ministry of Health as the most reliable source of information.^7^ These participants in our study were also found to have the correct information regarding the disease spread as per the Ministry of Health information.^8^ However, they were partially correct regarding the high-risk category of the patients who are older people and those with chronic diseases as opposed to children and adolescents, according to the WHO.^9 10^ Social media can be made widely available to the general population to raise and sustain awareness before, during and after lockdowns, regarding COVID-19; mechanisms by which it is transmitted and the precautionary measures which should be observed to minimize the disease transmission. The awareness campaigns can also include information regarding usual and unusual COVID-19 symptoms and how to manage them, along with long-term side effects of the disease and medications used to manage COVID-19. Further research can highlight whether good awareness can help prevent or curb the disease spread in society.

Our study participants appeared to be following the precautionary measures appropriately as per WHO recommendations.^11^ This can be linked to the good awareness and knowledge of COVID-19 which was reflected in the participants’ views. However, the participants highlighted that after the lifting of the lockdown, the precautionary measures were not being observed by the people in the same way as they were being observed during the lockdown period. This could be attributed to the false notion that the coronavirus is no longer dangerous since the lockdown has been lifted and, therefore, necessitates continuous awareness regarding the infection.

The initial symptoms reported by our study participants were in line with the initial symptoms reported by the patients in other studies.^12 13 14^ One of the participants experienced hair loss due to COVID-19. According to the American Academy of Dermatology Association, hair loss symptoms may be expected during COVID-19 illness as temporary hair loss is normal after a fever or acute illness.^15^

The severity of COVID-19 and disease progression have been shown to be directly linked to increased age and comorbidities.^16^ Poor COVID-19 outcomes have particularly been associated with cardiovascular-related comorbid conditions such as hypertension and diabetes.^17^ Some of our study participants with hypertension and diabetes also reiterated the worsening of their comorbid conditions as well as the COVID-19 illness, which could also be attributed to their increased age compared to the relatively younger participants. Comorbid conditions should be managed equally well especially in the elderly to prevent deterioration. The COVID-19 pandemic has had a direct impact on the medication supply worldwide.^18^ Since Nakkasah was under complete lockdown, there was a reduced supply of medications for other conditions, and this could have also contributed to the worsening of comorbid conditions, as highlighted by one of our study participants.

As COVID-19 is known to be highly contagious, the majority of the participants believed that when isolated or quarantined, they did not transmit the disease to others while they were infected. This again ensues from the fact that the majority of the participants also indicated good knowledge about how the disease spreads. One of the participants indicated that he/she caught the infection from his/her brother who was asymptomatic. Evidence suggests that the disease is believed to be mostly transmitted by people who are asymptomatic or in the pre-symptoms period.^19^

The majority of the participants reported using the medicines to control their symptoms. Some of them mentioned the use of antibiotics, which seems inappropriate to use in the COVID-19 infection as it is a viral infection. This can even be disadvantageous by increasing the bacterial resistance in the long run. However, in some cases, the use of antibiotics may be beneficial since COVID-19 sometimes co-exist with a bacterial infection according to the WHO.^20^ Others reported that the home remedies helped in their recovery from COVID-19. Since these complementary medicines have an anti-inflammatory effect, these might have played a role by increasing their immunity against COVID-19.^21^

The majority of the participants in Nakkasah live on daily wages, and thus the lockdown resulting in shut down of all work had a disastrous effect on their financial situation. This was reflected in some of our study participants’ narratives and has been a characteristic of lockdowns in this pandemic.^22^ Several long-term effects have been reported in COVID-19 patients after the recovery.^23^ Our participants also reported some of these lingering side effects, which could be attributed to COVID-19. However, these could also be the side effects of the medicines taken by them during their illness.

Support from family and friends can be key to the survival of individuals in any circumstances and it has been particularly highlighted in this pandemic.^24^ The majority of the participants in our study emphasized how the emotional support from family and friends had helped them during the illness. Similarly, the support from the authorities and volunteer organizations can be a key to the survival of the societies in any circumstances and it has been particularly highlighted in this pandemic.^25^ The role of the Saudi Ministry of Health and volunteer organizations during the lockdown was particularly acknowledged by our participants for taking care of their daily necessities.

This study had some limitations. One of the limitations was the translation from one language to the other at various stages of the study which could have affected the core meaning and expression. Firstly, the interview guide was prepared in the English language as the study supervisors were English-speaking. The research students, who were native Arabic speakers, then translated the interview guide into the Arabic language in which the interviews were conducted. Some participants could not speak the Arabic language and, therefore, the official translator had to be utilized for communicating with those participants. The interview transcripts were in the Arabic language, which we decided not to translate into the English language to maintain the true essence and expression of the language to some extent. However, the codes, subthemes and themes were developed in the English language to be reviewed with the supervisors. Only the relevant quotes from the interview transcripts, presented in the results section, were translated into the English language.

## CONCLUSION

This study presents the experiences of COVID-19 recovered patients from a hotspot during the lockdown. Social media can be made widely available to the general population to raise and sustain awareness before, during and after lockdowns, regarding COVID-19, mechanisms by which it is transmitted and the precautionary measures which should be observed to minimize the disease transmission. Comorbid conditions should be managed equally well especially in the elderly to prevent deterioration. Appropriate precautionary measures, including isolation, must always be observed during the COVID-19 illness to prevent the transmission to others. Mental health support should be considered for vulnerable patients. Families and friends should also consider financial support to the needy ones during lockdowns in addition to the emotional support. The authorities must ensure adequate food and medicines supply during the lockdowns. Our findings have implications for healthcare and volunteer organizations to tailor their care models and streamline the workflows for improving patients’ quality of life during and after their illness in high-risk communities, during this pandemic, and in future disease outbreaks.

### What is already known on this subject

We conducted a comprehensive literature search on 10 Dec 2020, for studies published in the English language in PubMed, EMBASE and Google Scholar. We found a few studies describing the experiences of COVID-19 recovered patients in which impact on mental health was the highlight, along with the emphasis on the symptoms. However, we could not find any studies that explored COVID-19 recovered patients’ experiences in a hotspot or under lockdown, which can significantly impact the individual experiences.

### What this study adds

Having suffered from COVID-19 while living under lockdown can significantly affect people’s lives in different aspects. Our study describes such people’s experiences and the lessons drawn from them. Social media can play a crucial role in enhancing awareness regarding coronavirus, symptoms of COVID-19 and the precautionary measures to take to prevent the disease transmission. Special attention must be paid to the simultaneous management of comorbid conditions and the mental health of these individuals. The authorities and volunteer organizations must ensure adequate food and medicines supply to these individuals during the lockdown.

The summary of our findings described above has implications for healthcare and volunteer organizations to tailor their care models and streamline the workflows for improving patients’ quality of life during and after their illness in high-risk communities, during this pandemic, and in future disease outbreaks.

## Data Availability

The authors are willing to share the interview transcripts (Arabic), the list of corresponding generated codes developed from these transcripts and the study protocol with the interested researchers involved in qualitative data synthesis. The data can be requested via email to the corresponding author.

## Acknowledgements

The authors acknowledge all the academic staff who helped in the translation and validation of the interview guide. The authors would also like to thank the community leader, the official translator of Nakkasah and all the participants for their cooperation throughout this study.

## Twitter

Abdulrahman Alhajjaji @i3bd8, Ahmad Kurdi @Ahmad0Kurdi, Sultan Faqeh @Sultan FH, Safwan Alansari @safwan_ansare, Akrm Abdulaziz @AkrmM7md, Moayad Allihyani @shrapek10, Omar Almaghamsi @omaralharbi1998, Ejaz Cheema @cheemae4, Majid Ali @maajidaali

## Contributors

MaA conceived and designed the study. MaA and EC supervised the study at every stage. AbA and OA conducted the interviews and double-checked the interview transcripts. AK, SF, SA, AkA and MoA transcribed the interviews. All authors contributed equally to the data analysis and writing, reviewing and editing of this manuscript. All authors have full access to all the data and take full responsibility for the accuracy of the information presented in this manuscript.

## Patient consent for publication

Not required.

## Provenance and peer review

Not commissioned; externally peer-reviewed.

## Data availability statement

Data available upon request. The authors are willing to share the interview transcripts (Arabic), the list of corresponding generated codes developed from these transcripts and the study protocol with the interested researchers involved in qualitative data synthesis. The data can be requested via email to the corresponding author.

---

**Panel 1.**
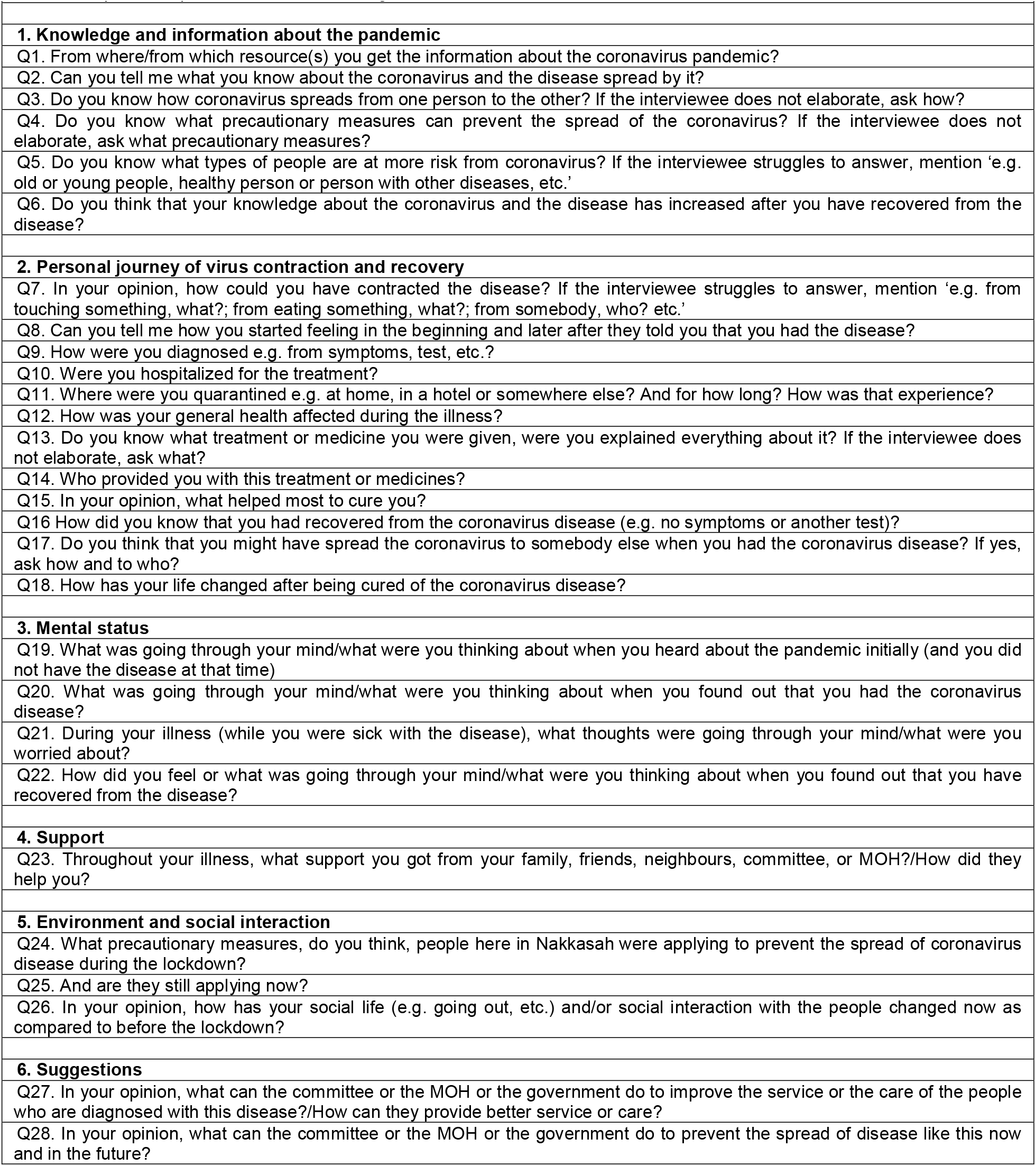
Topics and questions in the interview guide

**Panel 2.**
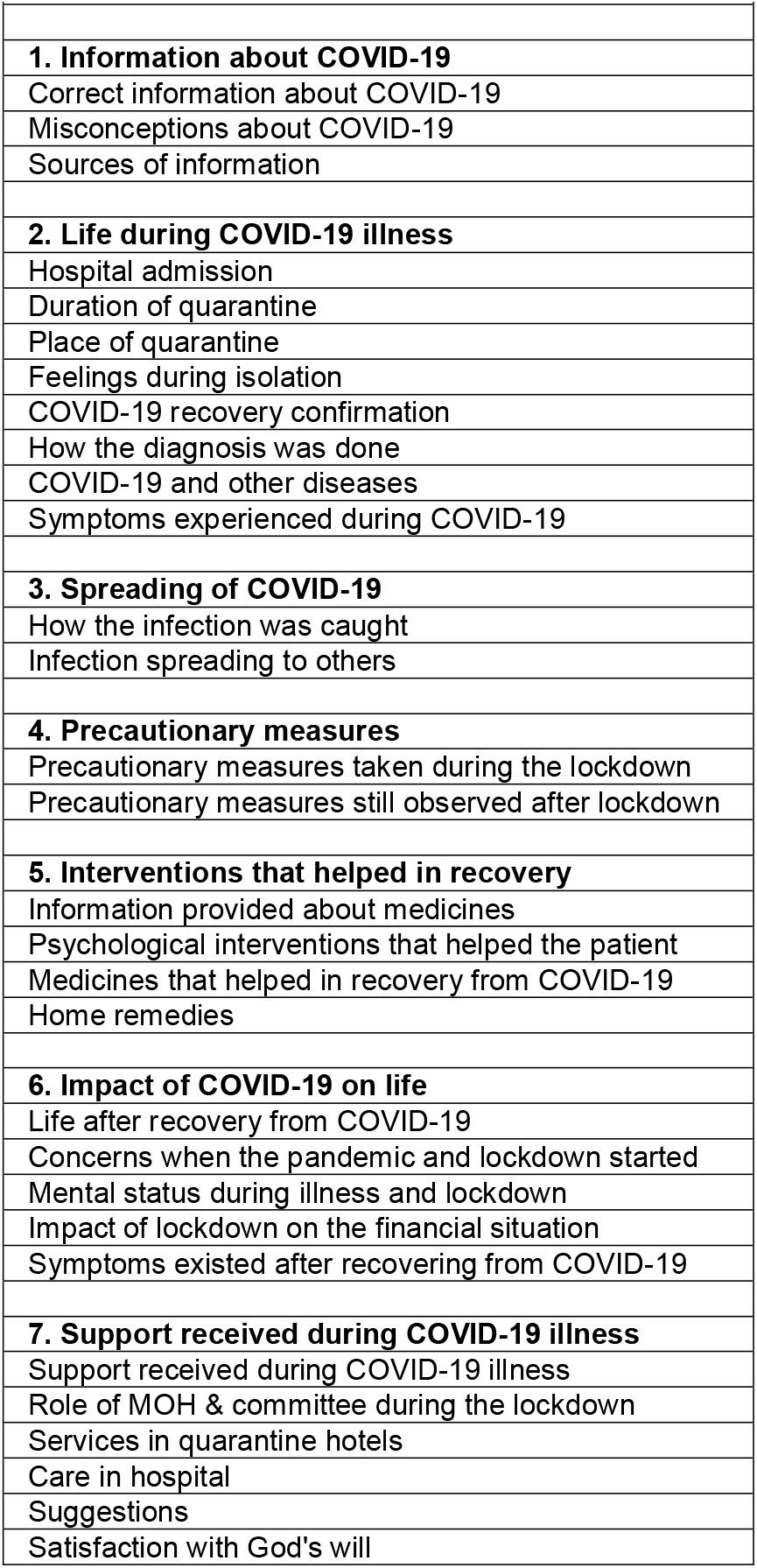
Themes and the associated subthemes

## REFERENCES

1. Rashed ER, Eissa ME. Global assessment of morbidity and mortality pattern of COVID-19: Descriptive statistics overview. Iberoam J Med 2020; 2: 68–72.

2. Kluge HHP. Older people are at highest risk from COVID-19, but all must act to prevent community spread. World Health Organization Regional Office for Europe, 2020. https://www.euro.who.int/en/health-topics/health-emergencies/coronavirus-covid-19/statements/statement-older-people-are-at-highest-risk-from-covid-19,-but-all-must-act-to-prevent-community-spread (accessed Jan 30, 2021).

3. Q&A. Coronavirus disease (COVID-19): How is it transmitted? World Health Organization, 2020. https://www.who.int/news-room/q-a-detail/coronavirus-disease-covid-19-how-is-it-transmitted (accessed Jan 30, 2021).

4. Abbas A. In Makkah, a new neighborhood with same old problems. Arab News, 2006. www.arabnews.com/node/284820 (accessed Jan 29, 2021).

5. Millstein JH, Kindt S. Reimagining the patient experience during the COVID-19 pandemic. NEJM Catalyst, 2020. https://catalyst.nejm.org/doi/full/10.1056/CAT.20.0349 (accessed Jan 30, 2021).

6. GMI Blogger. Saudi Arabia social media statistics 2020. Global Media Insight, 2020. https://www.globalmediainsight.com/blog/saudi-arabia-social-media-statistics/ (accessed Feb 3, 2021).

7. Alanezi F, Aljahdali A, Alyousef S, Alrashed H, Alshaikh W, Mushcab H, et al. Implications of public understanding of COVID-19 in Saudi Arabia for fostering effective communication through awareness framework. Frontiers in Public Health 2020; 8: 494. DOI: doi: 10.3389/fpubh.2020.00494

8. COVID-19 Awareness. How to protect yourself from COVID-19? Ministry of Health, 2020. https://covid19awareness.sa/en/home-page (accessed Feb 3, 2021).

9. UNICEF data. Child & adolescent health and COVID-19. United Nations International Children’s Emergency Fund, 2020. https://data.unicef.org/topic/child-health/child-health-and-covid-19/ (accessed Feb 3, 2021).

10. Information for the Public. COVID-19: vulnerable and high-risk groups. World Health Organization, 2020. https://www.who.int/westernpacific/emergencies/covid-19/information/high-risk-groups#:~:text=COVID-19 (accessed Feb 5, 2021).

11. Advice for the Public. Coronavirus disease (COVID-19) advice for the public. World Health Organization, 2021. https://www.who.int/emergencies/diseases/novel-coronavirus-2019/advice-for-public#:~:text= (accessed Feb 11, 2021).

12. Ge H, Wang X, Yuan X, et al. The epidemiology and clinical information about COVID-19. Eur J Clin Microbiol Infect Dis 2020; 39: 1011–9. DOI: 10.1007/s10096-020-03874-z.

13. Jiang F, Deng L, Zhang L, et al. Review of the Clinical Characteristics of Coronavirus Disease 2019 (COVID-19). J Gen Intern Med 2020; 35: 1545–9. DOI: 10.1007/s11606-020-05762-w.

14. Li H, Liu SM, Yu XH, Tang SL, Tang CK. Coronavirus disease 2019 (COVID-19): current status and future perspectives. Int J Antimicrob Agents 2020; 55: 105951. DOI: 10.1016/j.ijantimicag.2020.105951.

15. Diseases and Conditions. Can COVID-19 cause hair loss? American Academy of Dermatology Association, 2020. https://www.aad.org/public/diseases/hair-loss/causes/covid-19 (accessed Feb 5, 2021).

16. Yang J, Zheng YA, Gou X, et al. Prevalence of comorbidities and its effects in patients infected with SARS-CoV-2: A systematic review and meta-analysis. Int J Infect Dis 2020; 94: 91–5. DOI: 10.1016/j.ijid.2020.03.017

17. Sanyaolu A, Okorie C, Marinkovic A, et al. Comorbidity and its impact on patients with COVID-19. SN Compr Clin Med 2020; published online June 25. DOI:10.1007/s42399-020-00363-4

18. Gopalakrishnan BN, Vickers B, Ali S. Analysing the effects of the COVID-19 pandemic on medical supply chains in commonwealth countries. International Trade Working Paper, No. 2020/09, Commonwealth Secretariat, London, 2020. DOI: 10.14217/501dd683-en.

19. Moghadas SM, Fitzpatrick MC, Sah P, et al. The implications of silent transmission for the control of COVID-19 outbreaks. Proc Natl Acad Sci 2020; 117: 17513–5. DOI: 10.1073/pnas.2008373117.

20. Disease Prevention. Preventing the COVID-19 pandemic from causing an antibiotic resistance catastrophe. World Health Organization, 2020. https://www.euro.who.int/en/health-topics/disease-prevention/antimicrobial-resistance/news/news/2020/11/preventing-the-covid-19-pandemic-from-causing-an-antibiotic-resistance-catastrophe (accessed Feb 6, 2021).

21. Panyod S, Ho C-T, Sheen L-Y. Dietary therapy and herbal medicine for COVID-19 prevention: A review and perspective. J Tradit Complement Med 2020; 10: 420–7. DOI: 10.1016/j.jtcme.2020.05.004.

22. Carroll N, Sadowski A, Laila A, et al. The impact of COVID-19 on health behavior, stress, financial and food security among middle to high income Canadian families with young children. Nutrients 2020; 12: 2352. DOI: 10.3390/nu12082352

23. Lopez-Leon S, Wegman-Ostrosky T, Perelman C, et al. More than 50 long-term effects of COVID-19: A systematic review and meta-analysis. medRxiv 2021; published online Jan 30. DOI: 10.1101/2021.01.27.21250617.

24. Hart JL, Turnbull AE, Oppenheim IM, Courtright KR. Family-centered care during the COVID-19 era. J Pain Symptom Manage 2020; 60: e93–7. DOI: 10.1016/j.jpainsymman.2020.04.017

25. Miao Q, Schwarz S, Schwarz G. Responding to COVID-19: Community volunteerism and coproduction in China. World Dev 2021; 137: 105128. DOI: 10.1016/j.worlddev.2020.105128

